# Plasma Proteomic Classification of Alzheimer Disease, Parkinson Disease, and Amyotrophic Lateral Sclerosis Using a Single Neurodegeneration Panel: A Diagnostic Cohort Study

**DOI:** 10.64898/2026.10.19.26365782

**Authors:** Kensuke Okada, Sho Shimohama, Ryotaro Okochi, Shogyoku Bun, Keisuke Takahata, Hajime Tabuchi, Yu Iwabuchi, Toshiki Tezuka, Tomonori Nukariya, Shohei Okusa, Yoshihiro Nihei, Satoru Morimoto, Shinichi Takahashi, Kenji Tai, Yoshiaki Sato, Junro Kuromitsu, Jin Nakahara, Morinobu Seki, Hideyuki Okano, Masaru Mimura, Daisuke Ito

## Abstract

**Importance:** Blood biomarkers for Alzheimer disease(AD), Parkinson disease(PD), and amyotrophic lateral sclerosis(ALS) are typically developed as disease-specific single-analyte tests. Whether one shared plasma panel can support disease-selective classification remains unclear.

**Objective:** To test whether the Olink Target 48 Neurodegeneration plasma panel, analyzed with disease-specific Elastic Net models, can discriminate AD, PD, and ALS in a heterogeneous neurological cohort.

**Design, Setting, and Participants:** Single-center diagnostic cohort study at Keio University Hospital, Tokyo, Japan. After quality control and one consent withdrawal, 302 participants were analyzed(healthy controls n = 62; AD continuum n = 93; PD n = 39; ALS n = 27; other neurological disorders n = 81) and allocated by diagnosis-stratified sampling into a discovery training set(n = 211) and a locked hold-out test set(n = 91).

**Exposures:** Plasma normalized protein expression from 42 assays retained after protein quality control. Plasma DOPA decarboxylase was excluded a priori from all classifiers because blood levels primarily reflect dopaminergic treatment exposure.

**Main Outcomes and Measures:** Hold-out receiver operating characteristic area under the curve(AUC). Sensitivity and specificity at a training-set Youden threshold locked for Test.

**Results:** Hold-out AUCs were 0.974(95% CI, 0.942-0.995) for AD, 0.926(0.820-1.000) for PD, and 0.995(0.980-1.000) for ALS. At locked Youden cutoffs, sensitivity was 0.929 (95% CI, 0.765-0.991) and specificity was 0.921 (95% CI, 0.824-0.974) for AD, 0.833 (95% CI, 0.516-0.979) and 0.975 (95% CI, 0.912-0.997) for PD, and 1.000 (95% CI, 0.631-1.000) and 0.940 (95% CI, 0.865-0.980) for ALS.

**Conclusions and Relevance:** A single plasma neurodegeneration panel supported disease-selective classification of AD, PD, and ALS in internal validation. Multicenter external validation is needed before clinical application.

**Key Points:** *Question:* Can a single plasma neurodegeneration panel classify Alzheimer disease (AD), Parkinson disease (PD), and amyotrophic lateral sclerosis (ALS) in a heterogeneous neurological cohort?

*Findings:* In this diagnostic cohort study, disease-specific Elastic Net models derived from a single plasma panel enabled the discrimination of AD, PD, and ALS from other neurological disorders and healthy controls.

*Meaning:* A single plasma neurodegeneration panel can simultaneously support disease-selective classification of AD, PD, and ALS, offering a unified approach to blood-based diagnosis.

## Introduction

In Alzheimer disease (AD), plasma p-tau217 serves as a robust marker for established amyloid-beta pathology, demonstrating high diagnostic accuracy that often matches or exceeds cerebrospinal fluid measures ^1,2,3,4,5^. The plasma p-tau217/Aβ42 ratio reflects cerebral amyloid pathology and reduces borderline classifications compared to p-tau217 alone, while maintaining equivalent or superior discriminative ability ^6,7,8^. For Parkinson disease (PD), blood-based alpha-synuclein seed amplification assays (SAA) demonstrate high diagnostic accuracy and are positioned as high-sensitivity screening tools within a multimodal strategy, rather than standalone replacements for CSF alpha-synuclein SAA confirmation ^9,10^. In amyotrophic lateral sclerosis (ALS), blood neurofilament light chain demonstrates high diagnostic accuracy for distinguishing the disease from healthy controls and mimics, while also serving as a robust predictor of disease progression and survival ^11,12,13,14,15^.

Although useful single-analyte blood markers exist for individual diseases, a unified blood framework that distinguish between the major proteinopathies (AD, PD and ALS) together remains incomplete. Cross-cutting markers such as blood neurofilament light chain often reflect axonal injury intensity rather than disease-specific pathology. What is still missing is a shared blood platform on which these conditions can be evaluated side by side, rather than through separate single-analyte pipelines. Addressing this gap requires testing whether one accessible blood panel can support disease-versus-non-target discrimination across these conditions. Therefore, this study aims to demonstrate that a single blood panel combined with machine learning can discriminate AD, PD, and ALS in a heterogeneous neurological cohort.

## Methods

### Standard Protocol Approvals, Registrations, and Patient Consents

Institutional review board approval and written informed consent were obtained from all participants or their legally authorized representatives, where applicable. Approval numbers, trial registrations, and consent procedures are provided in the eMethods in Supplement 1.

### Study Design

This single-center observational diagnostic study was conducted at Keio University Hospital (Tokyo, Japan). Plasma proteins were measured with the Olink Target 48 Neurodegeneration panel (Olink Proteomics AB), reported as Normalized Protein eXpression (NPX), with 42 assays retained after protein quality control. The cohort comprised HC, AD, PD, ALS, and Others.

The analytical sample was assembled from three recruitment streams: a previously reported Japanese memory-clinic cohort contributing HC, AD, and Others ^16^; 20 ALS participants from ROPALS ^17^; and PD plus additional ALS participants from the Department of Neurology. Of 313 samples assayed, 11 were excluded (failed sample quality control, n = 5; median NPX outlier ≥3 SD, n = 1; missing data, n = 1; anxiety disorder, n = 3; consent withdrawal, n =1), leaving 302 participants (Table 1; Figure 1). Participants were allocated by diagnosis-stratified sampling (train ratio 70%) to a training (discovery) set (n = 211) and a prespecified hold-out test set (n = 91). Primary analyses were 3 disease-specific binary Elastic Net classifiers (AD vs non-AD, PD vs non-PD, and ALS vs non-ALS). Additional laboratory, preprocessing, and secondary statistical procedures are provided in the eMethods in Supplement 1.

**Figure 1.**
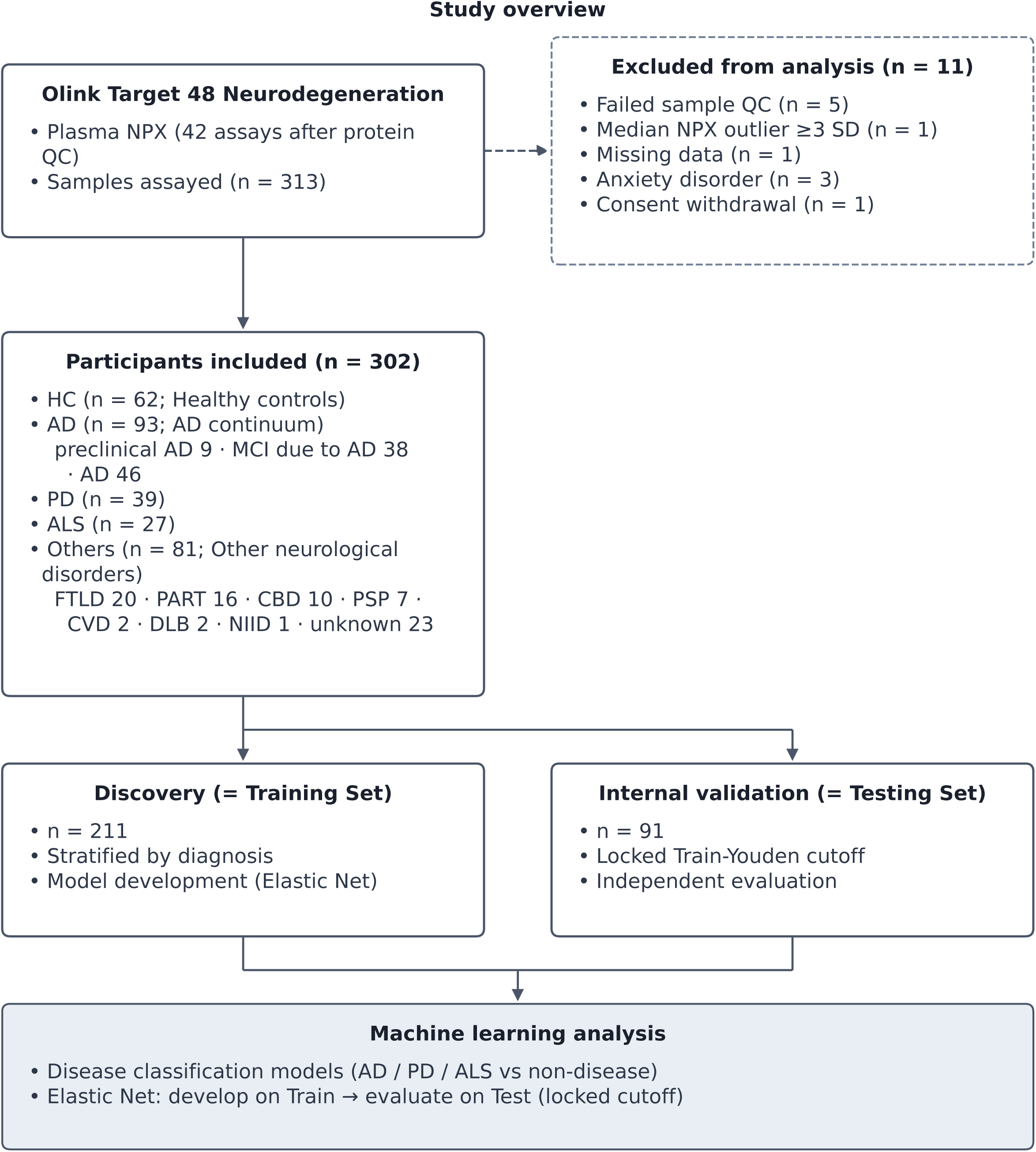
Study overview. Of 313 samples assayed using the Olink Target 48 Neurodegeneration panel (Plasma NPX; 42 assays retained after protein quality control), 11 were excluded (failed sample quality control, n=5; median NPX outlier ≥3 SD, n=1; missing data, n=1; anxiety disorder, n=3; and consent withdrawal, n=1), yielding an analytical cohort of n=302. The cohort comprised healthy controls (HC, n=62); the Alzheimer disease continuum (AD, n=93: preclinical AD, n=9; mild cognitive impairment due to AD, n=38; Alzheimer disease, n=46); Parkinson disease (PD, n=39); amyotrophic lateral sclerosis (ALS, n=27); and other neurological disorders (Others, n=81: frontotemporal lobar degeneration, n=20; suspected primary age-related tauopathy, n=16; corticobasal syndrome, n=10; progressive supranuclear palsy, n=7; cerebrovascular disease, n=2; dementia with Lewy bodies, n=2; neuronal intranuclear inclusion disease, n=1; and unknown etiology, n=23). Participants were allocated into a Discovery set (Training Set, n=211; diagnosis-stratified sampling; Elastic Net model development) and an Internal validation set (Testing Set, n=91; locked Train-Youden cutoff; independent evaluation). Machine learning analysis included disease classification models (AD, PD, and ALS versus non-disease) and Elastic Net models developed on the training set and evaluated on the test set using a locked cutoff.

**Table 1.** Baseline characteristics of the analytical cohort (n=302). Columns are healthy controls (HC); Alzheimer disease (AD) continuum; Parkinson disease (PD); amyotrophic lateral sclerosis (ALS); other neurological disorders (Others); and Total. The AD continuum comprises preclinical AD, mild cognitive impairment (MCI) due to AD, and AD. Others uses the paper reporting taxonomy (frontotemporal lobar degeneration [FTLD], primary age-related tauopathy [PART], corticobasal syndrome [CBS], progressive supranuclear palsy [PSP], cerebrovascular disease [CVD], dementia with Lewy bodies [DLB], neuronal intranuclear inclusion disease [NIID], and unknown; see Figure 1). Values are mean ± SD or n (%). Age is also summarized in bins (<50, 50-64, ≥65 years). Disease-specific clinical scores (Mini-Mental State Examination [MMSE], Clinical Dementia Rating [CDR], Hoehn and Yahr scale, Unified Parkinson’s Disease Rating Scale part III [UPDRS-III], levodopa equivalent daily dose [LEDD], ALS Functional Rating Scale-Revised [ALSFRS-R]) are shown only in groups in which they were ascertained.

| Characteristic | HC | AD | PD | ALS | Others | Total |
| --- | --- | --- | --- | --- | --- | --- |
| Sample size, n | 62 | 93 | 39 | 27 | 81 | 302 |
| Male, n (%) | 36 (58.1) | 47 (50.5) | 25 (64.1) | 18 (66.7) | 40 (49.4) | 166 (55.0) |
| Female, n (%) | 26 (41.9) | 46 (49.5) | 14 (35.9) | 9 (33.3) | 41 (50.6) | 136 (45.0) |
| Age, mean ± SD | 68.4 ± 7.9 | 72.1 ± 9.6 | 69.5 ± 10.9 | 64.0 ± 10.4 | 70.2 ± 9.9 | 69.7 ± 9.8 |
| <50, n (%) | 0 (0.0) | 2 (2.2) | 2 (5.1) | 2 (7.4) | 2 (2.5) | 8 (2.6) |
| 50–64, n (%) | 22 (35.5) | 19 (20.4) | 10 (25.6) | 12 (44.4) | 18 (22.2) | 81 (26.8) |
| ≥65, n (%) | 40 (64.5) | 72 (77.4) | 27 (69.2) | 13 (48.1) | 61 (75.3) | 213 (70.5) |
| MMSE, mean ± SD | 29.0 ± 1.1 | 23.1 ± 5.7 | 27.6 ± 2.4 | — | 24.9 ± 5.8 | 25.6 ± 5.2 |
| CDR, mean ± SD | 0.0 ± 0.0 | 0.5 ± 0.4 | — | — | 0.5 ± 0.5 | 0.4 ± 0.4 |
| Hoehn and Yahr scale, mean ± SD | — | — | 2.5 ± 0.8 | — | — | 2.5 ± 0.8 |
| UPDRS-III, mean ± SD | — | — | 33.7 ± 17.7 | — | — | 33.7 ± 17.7 |
| LEDD, mean ± SD | — | — | 423.3 ± 222.2 | — | — | 423.3 ± 222.2 |
| ALSFRS-R, mean ± SD | — | — | — | 39.3 ± 7.4 | — | 39.3 ± 7.4 |

### Participants

Participants were classified as HC, AD continuum, PD, ALS, or Others. In the dementia stream ^16,18^, amyloid PET-positive participants were assigned to the AD continuum (preclinical AD, mild cognitive impairment due to AD, or AD). Amyloid PET-negative participants were classified as HC or Others according to published clinical criteria. ALS diagnoses met Gold Coast criteria ^19^ except for the ROPALS subset ^17^. PD diagnoses met Movement Disorder Society clinical diagnostic criteria ^20^. Cognitive and motor scales (Mini-Mental State Examination, Clinical Dementia Rating global score, Hoehn and Yahr scale, MDS-UPDRS Part III, levodopa equivalent daily dose, and ALS Functional Rating Scale-Revised) were recorded where applicable. Baseline characteristics are shown in Table 1.

### Plasma Proteomic Assay

Venous blood was collected into EDTA tubes and processed on ice. Plasma was separated by centrifugation and stored at −80 °C. Proteins were quantified by proximity extension assay as NPX. DOPA decarboxylase (DDC) was excluded a priori from all classifiers because plasma DDC largely reflects dopaminergic treatment exposure rather than intrinsic disease biology ^21,22,23^. Biospecimen handling, assay quality control, and missing-data rules are detailed in the eMethods in Supplement 1.

### Statistical Analysis

Analyses used R, version 4.4.1. For each disease, an Elastic Net logistic model (α = 0.5) was trained to discriminate the target diagnosis from all remaining participants, using quality-control-retained NPX assays plus age and sex. The penalty parameter λ was selected by stratified nested cross-validation (5 outer × 5 inner folds) with receiver operating characteristic (ROC) area under the curve (AUC) as the tuning metric; the final λ was then chosen by stratified cross-validation on the full training partition and the model was refit on that partition. Discrimination on the locked hold-out test set was summarized by ROC AUC with percentile bootstrap 95% CIs. Sensitivity, specificity, and accuracy used a Youden index threshold maximized on training-set predicted probabilities and locked for test evaluation; a default probability cut point of 0.5 was not used. Correlated AUCs for the panel versus the disease-specific key marker (AD: pTau217; PD: EIF2AK2; ALS: NEFL) were compared with DeLong’s test. Reporting followed TRIPOD+AI ^24^. Cross-platform correlations, principal component analysis, heatmap clustering, one-way ANOVA, limma differential expression, stratified Spearman correlations, SHAP interpretation, calibration, and robustness analyses are described in the eMethods in Supplement 1.

### Standard Protocol Approvals, Registrations, and Patient Consents

This study was conducted in accordance with the Declaration of Helsinki. Written informed consent was obtained from all participants or their legally authorized representatives when required. The dementia-stream cohort (HC, AD, and Others) was approved by the Certified Review Board of Keio University (N20170237) and registered with the University Hospital Medical Information Network Clinical Trials Registry (UMIN-CTR; UMIN000032027) and the Japan Registry of Clinical Trials (jRCT; jRCTs031180225), as described previously ^16^. Twenty participants with ALS were enrolled in the ROPALS study under approval of the Institutional Review Board of Keio University Hospital (D18-01) and the Keio University School of Medicine Ethics Committee (20080016) ^17^. Participants with PD and the remaining ALS cases were enrolled under Certified Review Board of Keio University approval N20221112.

### Diagnostic criteria and clinical assessments

Participants were classified into five groups: HC, AD continuum, PD, ALS, and Others (frontotemporal lobar degeneration [FTLD], n = 20; suspected primary age-related tauopathy [PART], n = 16; corticobasal syndrome [CBS], n = 10; progressive supranuclear palsy [PSP], n = 7; cerebrovascular disease [CVD], n = 2; dementia with Lewy bodies [DLB], n = 2; neuronal intranuclear inclusion disease [NIID], n = 1; unknown, n = 23). In the dementia cohort ^16^, participants with a positive amyloid PET scan were classified into the AD continuum, comprising preclinical AD, mild cognitive impairment due to AD, and AD. Participants with a negative amyloid PET scan were classified as HC or Others according to established clinical diagnostic criteria; clinical diagnoses within Others followed published criteria for the corresponding disorders, including FTLD ^25^, PSP ^26^, CBS ^27^, and DLB ^28^. Participants who were cognitively normal or had amnesia, and who were [18F]florbetaben amyloid PET-negative but [18F]florzolotau tau PET-positive in the medial temporal lobe, were classified as suspected PART. Twenty participants with ALS were enrolled from the ROPALS clinical trial ^17^, and additional participants with ALS were recruited from the Department of Neurology, Keio University Hospital, and met the Gold Coast diagnostic criteria for ALS ^19^. Participants with PD were recruited from the Department of Neurology, Keio University Hospital, and met the MDS clinical diagnostic criteria for PD ^20^.

Cognitive performance was assessed with the Mini-Mental State Examination (MMSE) ^29^. Dementia severity was assessed with the Clinical Dementia Rating global score (CDR-global) ^30^. Motor and disease-severity measures were assessed where applicable: the Hoehn and Yahr scale ^31^, MDS-UPDRS Part III ^32^, and levodopa equivalent daily dose (LEDD) ^33^ in patients with PD; and the ALS Functional Rating Scale-Revised (ALSFRS-R) ^34^ in patients with ALS.

Demographic and clinical characteristics of the analytical cohort (n = 302; HC n = 62, AD n = 93, PD n = 39, ALS n = 27, Others n = 81) are summarized in Table 1; corresponding characteristics of the discovery training set (n = 211) and the internal validation testing set (n = 91) are shown in eTable 1 and eTable 2, respectively. Pairwise cross-platform correlations used complete cases for each analyte pair (eFigure 1; eFigure 2).

### Biospecimen handling and proteomic measurement

Venous blood was collected into BD Vacutainer K2EDTA tubes (Hemogard closure; 13 × 100 mm; 6 mL; catalog no. 365900; Becton, Dickinson and Company). Immediately after collection, tubes were placed on ice and centrifuged at 1,200 × *g* for 10 min at 4 °C. Plasma was transferred and centrifuged again at 2,800 × *g* for 10 min at 4 °C. Clarified plasma was aliquoted into Thermo Scientific Matrix 2D storage tubes (catalog no. 3742-BR; Thermo Fisher Scientific) and stored at −80 °C until analysis. Processing from venipuncture to frozen storage was completed within 3 h. For participants enrolled in the ROPALS (ropinirole) clinical trial, plasma was collected before treatment initiation, and collection followed the trial-specified blood-sampling protocol ^17^.

Proteins were measured using the Olink Target 48 Neurodegeneration panel (Olink Proteomics AB, Uppsala, Sweden). The Olink proximity extension assay uses pairs of oligonucleotide-labeled antibodies and reports relative abundance as Normalized Protein eXpression (NPX) on a log2 scale ^35^. Panel-specific measurement, normalization, and control procedures followed the manufacturer documentation ^36^.

Assay-level quality control followed LOD/2 substitution for values below the limit of detection and exclusion of proteins or samples with missingness above 0.5 (see Proteomic data preprocessing).

### Proteomic data preprocessing

Plasma proteomic measurements were obtained using the Olink Target 48 Neurodegeneration panel. Values below the manufacturer-defined limit of detection (LOD) for each assay were handled according to the Olink quality control protocol. Proteins with missingness above 0.5 and samples with missingness above 0.5 were excluded. Remaining missing values were imputed using median imputation. Continuous predictors were scaled using z-score standardization. Covariates (age, sex) were included in adjusted models where applicable.

Candidate predictors for the binary Elastic Net models were the Olink Neurodegeneration NPX assays retained after QC, together with the prespecified covariates age and sex, with one a priori exclusion: DOPA decarboxylase (DDC). Plasma DDC was omitted from the candidate set for all three disease-specific classifiers (AD versus non-AD, PD versus non-PD, and ALS versus non-ALS) because blood DDC primarily reflects dopaminergic treatment exposure (including levodopa equivalent daily dose, LEDD) rather than intrinsic disease biology, limiting its interpretability as a diagnostic classifier feature ^21,22,23^. Feature selection among the remaining candidates was performed by the Elastic Net penalty within nested cross-validation (no separate univariate filter).

### Statistical software

Statistical analyses were performed using R (version 4.4.1) ^37^. Penalized logistic regression used the tidymodels framework with glmnet ^38^; model performance metrics used yardstick ^39^. Differential expression analysis used limma where applicable ^40^. Figures were generated using ggplot2 (version 4.0.2) ^41^. A fixed analysis-wide random seed (42) was used for reproducibility. Procedures specific to Figures 2-4 and eFigures 1-5 are detailed below.

**Figure 2.**
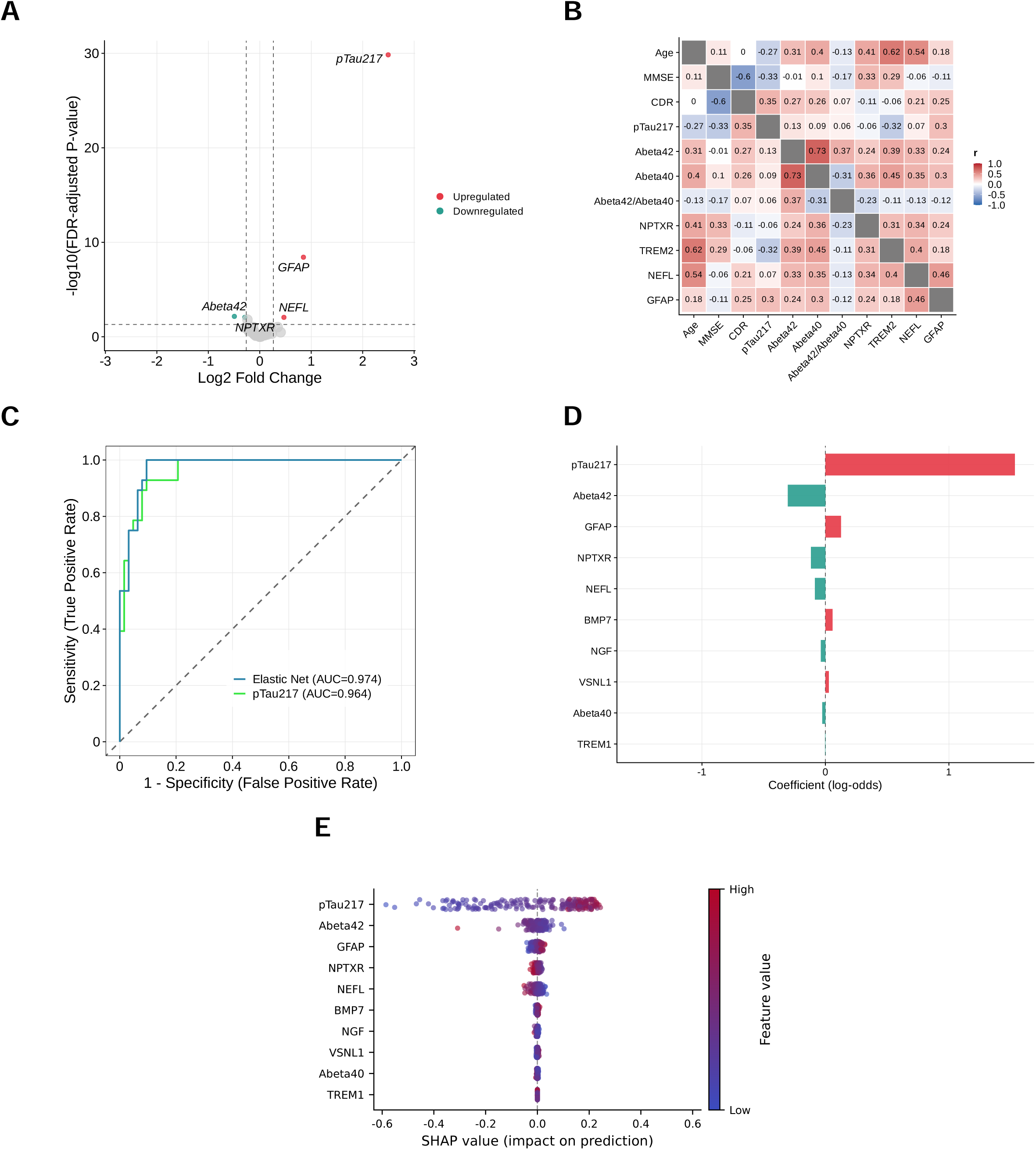
Plasma protein analyses in AD (A) Volcano plot of differential expression between AD and healthy controls (HC; FDR = 0.05; fold-change = 1.2). Upregulated proteins include pTau217, GFAP, and NEFL; downregulated proteins include Aβ42 and NPTXR. Points are colored red for upregulated and green for downregulated proteins. (B) Stratified Spearman correlation heatmap in AD (n = 93) for 11 variables: Age, MMSE, CDR, pTau217, Aβ42, Aβ40, Aβ42/Aβ40, NPTXR, TREM2, NEFL, and GFAP. Cell color denotes the Spearman ρ coefficient. (C) Receiver operating characteristic (ROC) curves for AD versus non-AD classification. The Elastic Net model (blue curve) used a threshold locked at the training-set Youden index of 0.365, yielding a test AUC of 0.974 (95% CI, 0.942-0.995), sensitivity of 0.929, and specificity of 0.921. At this threshold, 26 of 28 AD cases were classified correctly (2 missed), with 5 false positives among 63 non-AD participants. This is overlaid with the ROC curve for pTau217 alone (green curve; AUC = 0.964; 95% CI, 0.932-0.996). (D) Elastic Net signed coefficients for all non-zero terms, including age and sex as covariates. Bars are colored red for positive coefficients (associated with the case group), green for negative coefficients (associated with the control group), and gray for covariates. (E) Global SHAP beeswarm plot illustrating feature contributions to Elastic Net predictions. Each point represents one participant; the horizontal axis indicates the SHAP value (impact on model prediction), and point color encodes the feature value from low (blue) to high (red). Features are ordered by mean absolute SHAP value. Leading features include pTau217, Aβ42, GFAP, NEFL, and NPTXR.

**Figure 3.**
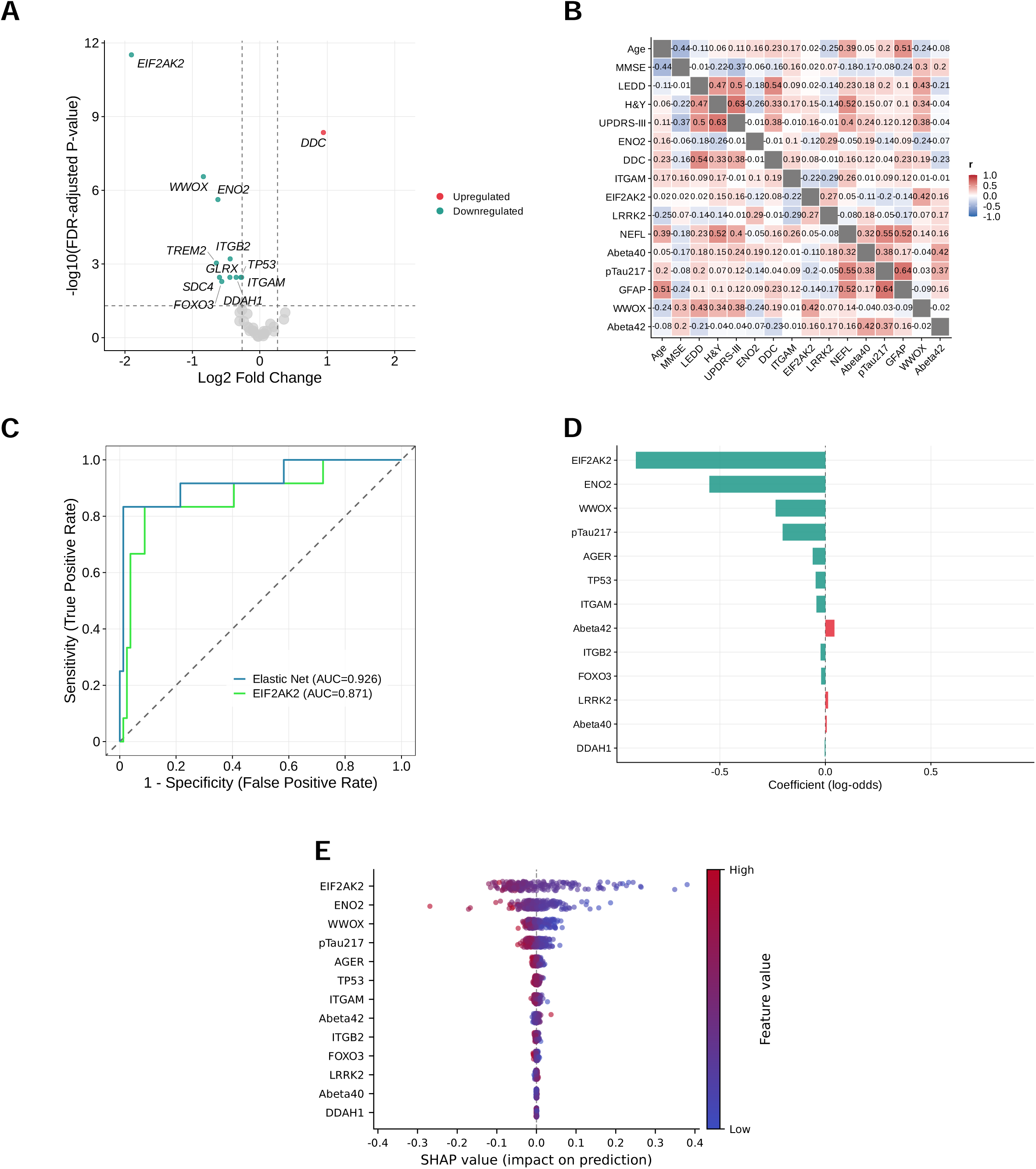
Plasma protein analyses in PD. (A) Volcano plot of differential expression between PD and healthy controls (HC; FDR = 0.05; fold-change = 1.2). DDC was upregulated, whereas EIF2AK2, WWOX, ENO2, ITGB2, TREM2, DDAH1, GLRX, ITGAM, SDC4, TP53, and FOXO3 were downregulated. Points are colored red for upregulated and green for downregulated proteins. (B) Stratified Spearman correlation heatmap in PD (n = 33) for 16 variables: Age, MMSE, LEDD, Hoehn and Yahr scale, UPDRS-III, ENO2, DDC, ITGAM, EIF2AK2, LRRK2, NEFL, Aβ40, pTau217, GFAP, WWOX, and Aβ42. Cell color denotes the Spearman ρ coefficient. (C) Receiver operating characteristic (ROC) curves for PD versus non-PD classification. The Elastic Net model (blue curve) used a threshold locked at the training-set Youden index (0.235), yielding a test AUC of 0.926 (95% CI, 0.820-1.000), with a test sensitivity of 0.833 and specificity of 0.975. At this threshold, 10 of 12 PD cases were classified correctly (2 missed), with 2 false positives among 79 non-PD participants. The curve for EIF2AK2 alone (green curve) is overlaid for comparison (AUC = 0.871; 95% CI, 0.744-0.999). (D) Elastic Net signed coefficients for all non-zero terms, including age and sex as covariates. Bars are colored red for positive coefficients (case), green for negative coefficients (control), and gray for covariates. (E) Global SHAP beeswarm plot illustrating feature contributions to Elastic Net predictions. Each point represents one participant; the horizontal axis indicates the SHAP value (impact on model prediction), and point color encodes the feature value from low (blue) to high (red). Features are ordered by mean absolute SHAP value. Leading features include EIF2AK2, ENO2, WWOX, pTau217, and AGER.

**Figure 4.**
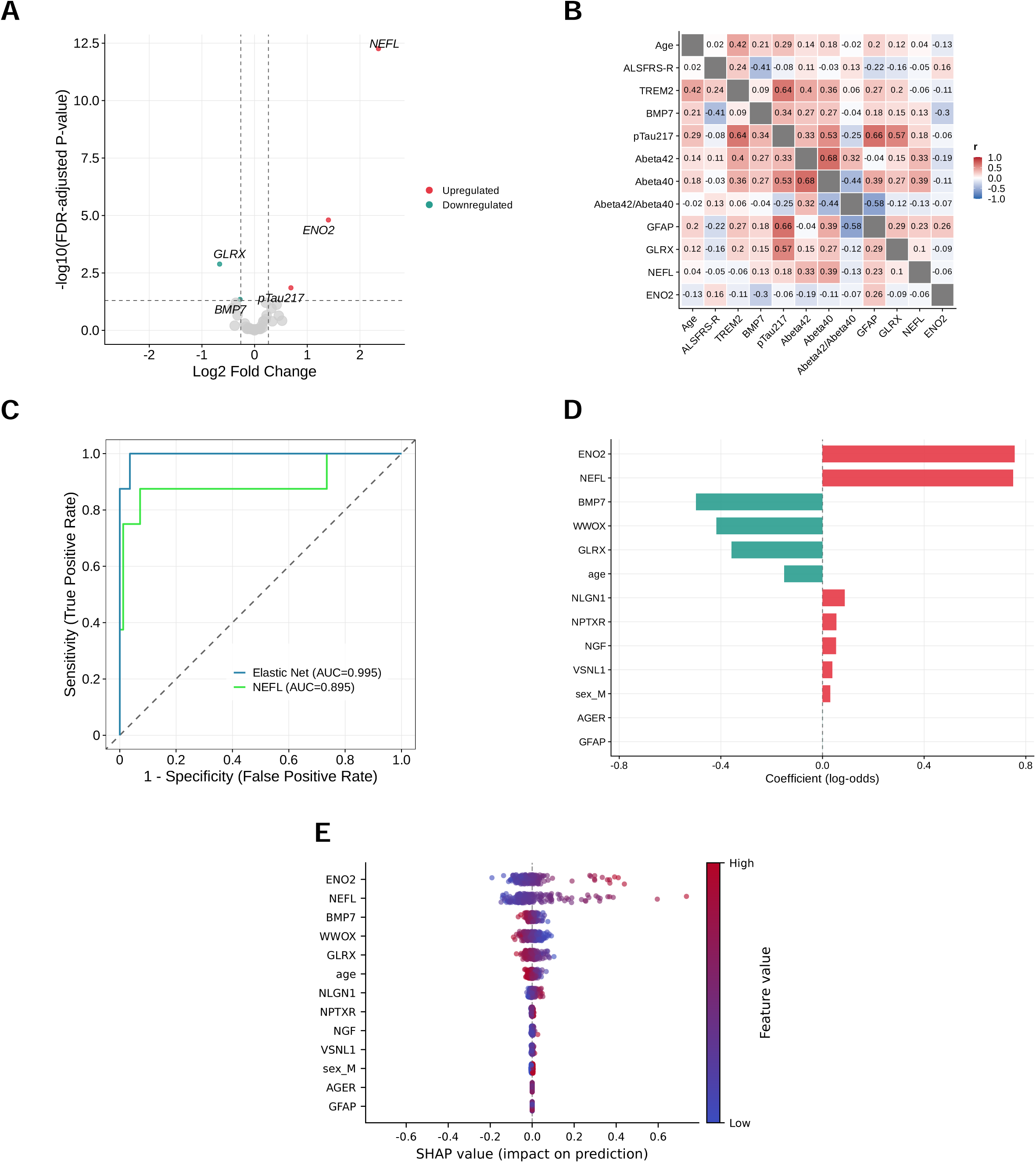
Plasma protein analyses in ALS. (A) Volcano plot of differential expression between ALS and healthy controls (HC; FDR = 0.05; fold-change = 1.2). Upregulated proteins include NEFL, ENO2, and pTau217; downregulated proteins include GLRX and BMP7. Points are colored red for upregulated and green for downregulated proteins. (B) Stratified Spearman correlation heatmap in the ALS cohort (n = 27) for 12 variables: Age, ALSFRS-R, TREM2, BMP7, pTau217, Aβ42, Aβ40, Aβ42/Aβ40, GFAP, GLRX, NEFL, and ENO2. Cell color denotes the Spearman ρ coefficient. (C) Receiver operating characteristic (ROC) curves for distinguishing ALS from non-ALS cases. The Elastic Net model (blue curve) used a threshold locked at the training-set Youden index of 0.090, yielding a test AUC of 0.995 (95% CI, 0.980-1.000), with a test sensitivity of 1.000 and specificity of 0.940. At this threshold, all 8 ALS cases were classified correctly, with 5 false positives among 83 non-ALS participants. The performance of NEFL alone is overlaid for comparison (green curve; AUC = 0.895; 95% CI, 0.717-1.000). (D) Elastic Net signed coefficients for all non-zero terms, including age and sex as covariates. Bars are colored red for positive coefficients (associated with the case group), green for negative coefficients (associated with the control group), and gray for covariates. (E) Global SHAP beeswarm plot illustrating feature contributions to Elastic Net predictions. Each point represents one participant; the horizontal axis indicates the SHAP value (impact on model prediction), and point color encodes the feature value from low (blue) to high (red). Features are ordered by mean absolute SHAP value. Leading features include NEFL, ENO2, WWOX, GLRX, and age.

### Cross-platform correlation

Pearson product-moment correlations were computed on the full analytical cohort between Olink Neurodegeneration NPX and matched values from orthogonal plasma platforms: SIMOA (pTau217, GFAP, and NEFL), HISCL (Aβ40, Aβ42, and the Aβ42/Aβ40 assay ratio), and High-Throughput (HT) assays (35 matched analytes). For display, ordinary-least-squares regression lines with 95% confidence bands were fitted; these fits were not used as classifier inputs.

### Principal component analysis

Principal component analysis (PCA) was performed on z-scored NPX values for the 42 proteins retained after protein quality control in the analytical cohort (n = 302). The first three principal components were examined.

### Proteomic profile heatmap

A heatmap of z-scored NPX values was generated for 12 core assays in the analytical cohort (n=302). Samples were annotated by diagnostic group. Hierarchical clustering used Ward linkage with Euclidean distance. The 12 assays were Aβ40, Aβ42, DDC, VSNL1, TREM2, GFAP, pTau217, EIF2AK2, NEFL, ENO2, S100B, and LRRK2.

### One-way ANOVA

Differences among diagnostic groups for each of the 42 proteins were assessed using one-way analysis of variance (ANOVA). Post-hoc pairwise comparisons used Tukey’s Honest Significant Difference (HSD) test. Multiple testing across proteins was summarized with Benjamini-Hochberg-adjusted ANOVA *P* values ^42^. *P*-value < 0.05 was considered statistically significant unless otherwise noted.

### Differential expression analysis

Differential expression analysis (limma) comparing each disease versus HC was performed on the discovery (training) cohort using linear models with empirical-Bayes variance moderation ^40^. Proteins with adjusted P < 0.05 and absolute fold change ≥ 1.2 (applied as |log2 fold change| ≥ log2(1.2)) were considered differentially expressed. Multiple testing was controlled using the Benjamini-Hochberg false-discovery-rate procedure ^42^.

### Exploratory correlation analysis

As an exploratory analysis, Spearman rank correlations were computed within disease strata for the following locked variable sets (cell-wise *P* values were not corrected for multiple comparisons):

- AD (n = 93, 11 variables): Age, MMSE, CDR, pTau217, Aβ42, Aβ40, Aβ42/Aβ40, NPTXR, TREM2, NEFL, GFAP
- PD (n = 33 complete cases of 39 PD participants, 16 variables): Age, MMSE, LEDD, Hoehn and Yahr scale, MDS-UPDRS Part III, ENO2, DDC, ITGAM, EIF2AK2, LRRK2, NEFL, Aβ40, pTau217, GFAP, WWOX, Aβ42
- ALS (n = 27, 12 variables): Age, ALSFRS-R, TREM2, BMP7, pTau217, Aβ42, Aβ40, Aβ42/Aβ40, GFAP, GLRX, NEFL, ENO2

### Predictive modeling

Three disease-specific elastic-net logistic models (α = 0.5) were developed on the training set to discriminate AD versus non-AD, PD versus non-PD, and ALS versus non-ALS. For each model, the positive class was the target disease and the negative class comprised all remaining participants. LOD/2 substitution, missingness filtering, median imputation, and standardization were applied to the training and hold-out partitions. Within tidymodels, categorical predictors were dummy encoded and numeric predictors were median-imputed, standardized, and screened for zero variance within each resampling analysis. The L1/L2 penalty (lambda) was tuned by stratified nested cross-validation (5 outer × 5 inner folds) over a regular grid of 20 lambda values from 1e-05 to 10, with ROC AUC as the selection metric. Nested cross-validation provided an internal estimate of discrimination. The final lambda was then selected by a further stratified cross-validation on the complete training partition, and the model was refit on that partition. Modeling used glmnet ^38^ and was reported with reference to TRIPOD+AI ^24^.

Predicted class probabilities were obtained from the fitted logistic model:

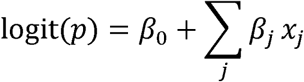

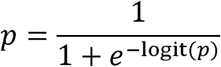

where x□denotes the preprocessed predictors retained with non-zero Elastic Net coefficients (continuous inputs were z-score standardized). Discrimination was summarized by ROC AUC, with a 95% confidence interval from percentile bootstrap resampling of hold-out predictions (2000 resamples) and a label-permutation test of ROC AUC (1000 permutations). Differences between correlated ROC AUCs for the panel model and the corresponding single key-marker model on the hold-out set were assessed using DeLong’s test. Model complexity was summarized by the events-per-variable ratio (minority-class size divided by the number of estimated parameters). Classification operating points followed the Youden threshold rule described below.

### Classification threshold

For primary reporting of sensitivity, specificity, and accuracy on the hold-out test set, the classification threshold was selected by maximizing Youden’s index (J = sensitivity + specificity − 1) ^43^ on the training-set predicted probabilities and then locked for Test evaluation. A default probability cutoff of 0.5 was not used for the main operating-point metrics. ROC AUC remains threshold-free. The threshold was not re-optimized on the hold-out test set.

Three disease-specific binary Elastic Net models were evaluated under this shared rule (Youden threshold from training, locked for Test):

- AD versus non-AD: Youden cutoff 0.365 (Test: 28 AD, 63 non-AD).
- PD versus non-PD: Youden cutoff 0.235 (Test: 12 PD, 79 non-PD).
- ALS versus non-ALS: Youden cutoff 0.090 (Test: 8 ALS, 83 non-ALS).

### Hold-out validation

Each fitted model was applied once to the pre-specified hold-out test set without refitting, feature reselection, or hyperparameter re-tuning. Operating-point metrics used the training-derived Youden threshold without re-optimization on the test set. Calibration was assessed graphically on the hold-out set by comparing observed event proportions with mean predicted probabilities in probability bins (45-degree line = perfect calibration), and quantified by calibration slope, calibration-in-the-large (intercept), and the Brier score.

### Model interpretation

SHapley Additive exPlanations (SHAP) were estimated for each fitted Elastic Net model using the fastshap Monte Carlo approximation (50 simulations per prediction) ^44^. The prediction function returned the probability of the prespecified positive class. The preprocessed predictor matrix from the 211 training observations served as the reference distribution. Global feature importance was summarized as the mean absolute SHAP value across observations; signed per-observation values were retained for local interpretation.

### Sensitivity and robustness analyses

Sensitivity analyses included (i) single-biomarker logistic models using the disease-specific key marker (AD: pTau217; PD: EIF2AK2; ALS: NEFL), (ii) multivariable Elastic Net models excluding that key marker, and (iii) leave-one-subgroup-out evaluations where subgroup labels were available. These secondary analyses were distinct from the primary hold-out classification analysis.

## Results

### Cohort characteristics

The analytical cohort comprised 302 participants (HC n = 62, AD continuum n = 93, PD n = 39, ALS n = 27, Others n = 81; Table 1). Mean age was 69.7 ± 9.8 years, with 166 participants (55.0%) male. Age ranged from 72.1 ± 9.6 years in AD continuum to 64.0 ± 10.4 years in ALS. Cognitive scores varied by diagnosis: MMSE was 29.0 ± 1.1 in HC versus 23.1 ± 5.7 in AD, where CDR-global was 0.5 ± 0.4. In PD, Hoehn and Yahr scale was 2.5 ± 0.8, MDS-UPDRS Part III was 33.7 ± 17.7, and LEDD was 423.3 ± 222.2. ALSFRS-R was 39.3 ± 7.4 in ALS. Baseline characteristics for the discovery training set (n = 211) and hold-out test set (n = 91) are detailed in eTable 1 and eTable 2.

### Study overview and analytical validation

We described cohort assembly and model development splits. From 313 samples assayed on the Olink Target 48 Neurodegeneration panel (plasma NPX; 42 assays retained after quality control), 11 were excluded (failed quality control, n = 5; median NPX outlier ≥3 SD, n = 1; missing data, n = 1; anxiety disorder, n = 3; consent withdrawal, n = 1), yielding an analytical cohort of 302 participants (Figure 1). The AD continuum included preclinical AD (n = 9), mild cognitive impairment due to AD (n = 38), and AD (n = 46). Other groups comprised FTLD (n = 20), PART (n = 16), CBD (n = 10), PSP (n = 7), CVD (n = 2), DLB (n = 2), NIID (n = 1), and unknown (n = 23). Participants were allocated via diagnosis-stratified sampling into a discovery training set (n = 211) and a pre-specified hold-out test set (n = 91) for disease-specific binary Elastic Net classifiers (AD, PD, and ALS versus non-target diagnoses). Operating-point metrics used a Youden threshold determined on the training set and locked for the hold-out test set.

Analytic validity was assessed by correlating Olink NPX with orthogonal plasma assays in the full cohort (eFigure 1). Pearson correlations were strong for SIMOA pTau217 (r = 0.919; 95% CI, 0.897-0.937; n = 238), GFAP (r = 0.898; 95% CI, 0.871-0.920; n = 238), and NEFL (r = 0.876; 95% CI, 0.842-0.903; n = 234), and for HISCL Aβ40 (r = 0.834; 95% CI, 0.791-0.869; n = 237). Correlations were more modest for HISCL Aβ42 (r = 0.467; 95% CI, 0.361-0.561; n = 238) and the Aβ42/Aβ40 assay ratio (r = 0.366; 95% CI, 0.251-0.471; n = 238). Concordance with High-Throughput assays for 35 matched analytes is shown in eFigure 2. While correlation between Olink Explore HT and the Target 48 Neurodegeneration Plasma Panel is generally high, weaker correlation were observed for FOXO3, STX1B, TP53, and WWOX (eFigure 2).

Global proteomic structure was inspected next. Principal-component analysis of the 42 scaled proteins (n = 302) indicated that the first three components explained 17.2%, 8.0%, and 6.4% of variance, respectively (cumulative 31.5%; eFigure 3). Diagnostic groups occupied overlapping regions of PC space rather than forming fully separated clusters, indicating that the proteomic variation comprises both shared and disease-specific components. A clustered heatmap of 12 core assays (including Aβ40, Aβ42, pTau217, GFAP, NEFL, DDC, EIF2AK2, ENO2, TREM2, VSNL1, S100B, and LRRK2) summarized group-annotated NPX profiles (eFigure 4). One-way ANOVA across diagnostic groups for the 42 proteins, with Tukey post-hoc pairwise contrasts, highlighted proteins with multi-group differences (eFigure 5; panels ordered by ascending Benjamini-Hochberg-adjusted ANOVA *P*; the derived Aβ42/Aβ40 ratio was excluded).

### Plasma proteomic profiling and classification of the AD continuum

#### Disease-associated plasma protein features

In AD versus HC, five proteins met FDR < 0.05 and |fold-change| ≥ 1.2 (Figure 2A). The strongest elevation was pTau217 (log2 fold-change = 2.495; adjusted *P* = 1.45×10□³□), followed by GFAP (0.848; 3.78×10□□) and NEFL (0.469; 8.74×10□³). Aβ42 (−0.494; 6.88×10□³) and NPTXR (−0.292; 8.74×10□³) were lower in AD.

Within AD continuum (n=93), exploratory Spearman correlations among 11 pre-specified variables were examined (Figure 2B). Higher pTau217 correlated with worse cognition and staging (MMSE ρ = −0.328, *P* = 0.0013; CDR ρ = 0.351, *P* = 0.0006). MMSE also correlated positively with NPTXR (ρ = 0.328, *P* = 0.0013). Among proteins, Aβ42 and Aβ40 were strongly correlated (ρ = 0.727, *P* < 0.001), and NEFL correlated with GFAP (ρ = 0.464, *P* = 3.7×10□□). Age correlated with TREM2 (ρ = 0.620, *P* = 3.3×10□¹¹) and NEFL (ρ = 0.539, *P* = 2.5×10□□). These cell-wise *P* values were not corrected for multiple comparisons.

#### Machine learning classification

A binary Elastic Net classifier discriminated AD from non-AD participants on the locked hold-out test set. The model achieved a Test AUC of 0.974 (95% CI, 0.942-0.995; Test: 28 AD, 63 non-AD) (Figure 2C). At the Youden threshold determined on the training set and locked for Test (0.365), sensitivity and specificity were 0.929 and 0.921, respectively. At this locked threshold, 26 of 28 AD cases were classified correctly (2 missed), with 5 false positives among 63 non-AD participants. A single-marker pTau217 model on the same hold-out partition yielded AUC 0.964 (95% CI, 0.932-0.996). The panel AUC was numerically higher than pTau217 alone, but DeLong’s test of correlated ROC curves did not show a significant difference (*P* = 0.35).

Non-zero model terms included pTau217, Aβ42, GFAP, NPTXR, NEFL, and BMP7 (Figure 2D). Global SHAP rankings likewise highlighted pTau217 together with Aβ42, GFAP, NEFL, and NPTXR (Figure 2E), aligning with the differential-expression pattern of higher pTau217/GFAP/NEFL and lower Aβ42/NPTXR in AD.

Secondary analyses assessed calibration and key-marker dependence. On the hold-out set (n=91), calibration metrics were Brier score 0.071, calibration slope 2.18, and calibration in the large 0.001. Excluding pTau217 from the multivariable Elastic Net reduced test AUC to 0.880 (sensitivity 0.857, specificity 0.730; ΔAUC −0.094 versus the robustness baseline). Leave-one-subgroup-out refits (excluding HC, Others, ALS, or PD from the training pool) retained high discrimination (test AUC 0.964-0.997). These analyses indicated that AD discrimination was strong and pTau217-centered: the panel did not significantly outperform pTau217 alone by DeLong’s test, whereas removing pTau217 clearly reduced performance.

### Plasma proteomic profiling and classification of PD

#### Disease-associated plasma protein features

Twelve proteins met FDR < 0.05 and |fold-change| ≥ 1.2 in PD versus HC (Figure 3A). DDC was higher in PD (log2 fold-change = 0.943; adjusted *P* = 4.45×10□□), whereas the largest decrease was EIF2AK2 (−1.903; 3.06×10□¹²), followed by WWOX (−0.837), ENO2 (−0.621), and other downregulated proteins. In complete-case PD participants (n = 33 of 39), exploratory Spearman correlations among 16 pre-specified variables were examined (Figure 3B). Motor severity measures were intercorrelated (Hoehn and Yahr scale with MDS-UPDRS Part III, ρ = 0.632, *P* = 0.0001). LEDD correlated positively with DDC (ρ = 0.542, *P* = 0.0011) and motor scores. Hoehn and Yahr scale also correlated with NEFL (ρ = 0.516, *P* = 0.0021). Among proteins, pTau217 correlated with GFAP (ρ = 0.644, *P* = 0.0001) and NEFL (ρ = 0.553, *P* = 0.0010). Cell-wise *P* values were uncorrected. Consistent with prior evidence that plasma DDC tracks dopaminergic therapy rather than disease biology ^21,22^, DDC was excluded a priori from candidate predictors for all three Elastic Net classifiers.

#### Machine learning classification

A PD versus non-PD Elastic Net classifier was trained and evaluated with a training-derived Youden threshold locked for Test. The model achieved a hold-out Test AUC of 0.926 (95% CI, 0.820-1.000; Test: 12 PD, 79 non-PD) (Figure 3C). At the locked threshold (0.235), sensitivity was 0.833 and specificity 0.975. Ten of 12 PD cases were classified correctly, with 2 false positives among 79 non-PD participants. A single-marker EIF2AK2 model yielded AUC 0.871 (95% CI, 0.744-0.999). The panel AUC was higher than EIF2AK2 alone, and DeLong’s test of correlated ROC curves was significant (*P* = 0.016).

Non-zero model terms included EIF2AK2, ENO2, WWOX, pTau217, AGER, and TP53 (Figure 3D). Global SHAP rankings highlighted EIF2AK2, ENO2, WWOX, and pTau217 among leading contributors (Figure 3E), consistent with prominent EIF2AK2 downregulation. Secondary checks quantified calibration and robustness. Hold-out calibration metrics were Brier score 0.066, calibration slope 2.42, and calibration in the large 0.157. Excluding EIF2AK2 changed test AUC to 0.968 (ΔAUC +0.042; sensitivity 0.750, specificity 0.962). Leave-one-subgroup-out AUCs ranged from 0.902 to 0.981; excluding Others yielded the highest AUC (0.981) but reduced sensitivity to 0.667. These analyses suggested a broader multi-marker PD signature, with the panel outperforming EIF2AK2 alone by DeLong’s test and discrimination remaining good after dropping EIF2AK2.

### Plasma proteomic profiling and classification of ALS

#### Disease-associated plasma protein features

In ALS versus healthy controls, five proteins met FDR < 0.05 and |fold-change| ≥ 1.2 (Figure 4A). NEFL showed the largest elevation (log2 fold-change = 2.352; adjusted *P* = 5.49×10□¹³), followed by ENO2 (1.402; 1.57×10□□) and pTau217 (0.689; 0.014). GLRX (−0.664; 0.0013) and BMP7 (−0.275; 0.045) were lower in ALS.

Exploratory Spearman correlations among 12 pre-specified variables in ALS (n = 27) revealed an inverse correlation between ALSFRS-R and BMP7 (ρ = −0.410, *P* = 0.034) (Figure 4B). Stronger associations occurred among proteins, including pTau217 with GFAP (ρ = 0.662, *P* = 0.0002), TREM2 (ρ = 0.636, *P* = 0.0005), and GLRX (ρ = 0.570, *P* = 0.0023). Age correlated with TREM2 (ρ = 0.419, *P* = 0.030). Cell-wise *P* values were uncorrected; several clinical-proteomic pairs did not reach *P* < 0.05 in this modest sample.

#### Machine learning classification

An Elastic Net classifier discriminated ALS from non-ALS participants with a hold-out Test AUC of 0.995 (95% CI, 0.980-1.000; Test: 8 ALS, 83 non-ALS) (Figure 4C). At the locked Youden threshold (0.090), sensitivity was 1.000 and specificity 0.940, correctly classifying all 8 ALS cases with 5 false positives. A single-marker NEFL model yielded AUC 0.895 (95% CI, 0.717-1.000). Although the panel AUC was numerically higher, DeLong’s test was not significant (*P* = 0.27), consistent with the small ALS test sample.

Non-zero model terms included ENO2, NEFL, BMP7, WWOX, GLRX, and NLGN1 (Figure 4D). Global SHAP rankings highlighted NEFL, ENO2, WWOX, GLRX, and age (Figure 4E), matching the differential-expression signature.

Calibration metrics included a Brier score of 0.023, slope of 3.28, and calibration in the large of −0.040. Excluding NEFL reduced test AUC to 0.958 (ΔAUC −0.038; sensitivity 0.875, specificity 0.916). Leave-one-subgroup-out refits retained near-ceiling discrimination (test AUC 0.994-0.996). These analyses indicated an ALS signature led by axonal injury markers; while NEFL exclusion cost appreciable discrimination, the numerical gain of the panel over NEFL alone was not statistically significant by DeLong’s test.

## Discussion

In this study, disease-specific binary Elastic Net models built from a single Olink Target 48 Neurodegeneration plasma panel achieved strong hold-out discrimination for the AD continuum, PD, and ALS (Test AUCs 0.974, 0.926, and 0.995, respectively). Feature contributions differed by disease, centering on pTau217 and related AD markers, an EIF2AK2-led PD signature, and axonal-injury markers in ALS. This indicates that one shared assay panel can support multiple disease-selective classification tasks. Orthogonal plasma assays agreed closely with Olink for pTau217, GFAP, NEFL, and Aβ40 (eFigure 1). Agreement for Aβ42 and the Aβ42/Aβ40 ratio was weaker. Given that our previous study found that the measurement of HISCL Aβ40 and Aβ42 demonstrated extremely high performance^16^, amyloid terms on this panel need further analytical work before they are given the same weight as pTau217 or GFAP.

Plasma pTau217 is a strong Alzheimer disease blood marker ^1,3,45,46^ and dominated the Alzheimer classifier. GFAP was also higher in AD, consistent with an astroglial signal ^47^ (eFigure 4 and eFigure 5). While the multi-marker model was numerically better than pTau217 alone, the practical advantage is that one multiplex run can carry the AD, PD, and ALS models simultaneously.

Parkinson disease blood work has centered on α-synuclein, including seed-amplification assays ^9,10^. These assays are technically cumbersome and unsuitable for multiple-panel testing; thus, this panel does not include direct assessment of α-synuclein pathology. DOPA decarboxylase was higher in PD and tracked levodopa equivalent daily dose, so we excluded it as a treatment-exposure marker ^21,22,23^. After exclusion, the leading PD signals were lower plasma EIF2AK2 and WWOX. EIF2AK2 (protein kinase R) phosphorylates eIF2α in the integrated stress response, while WWOX is an oxidoreductase implicated in neuronal maintenance. These are novel PD blood signals. A future panel adding α-synuclein assessment could test PD against other parkinsonian disorders, including multiple system atrophy, which was not in the present contrast group.

NEFL is the established ALS blood marker of axonal injury ^12,13^. Although NEFL is not disease-specific, as it reflects axonal injury across various neurodegenerative diseases, it remains a valuable marker for assessing disease severity and progression ^48^. In our model, NEFL was the leading feature, consistent with its role as a robust surrogate for motor neuron injury. However, the model also incorporated ENO2, BMP7, and WWOX, which were not prominent in the AD or PD classifiers. This suggests that the panel captures a disease-specific signature beyond the shared axonal-injury signal. By integrating NEFL with these additional proteins, the ALS model enhanced discrimination against other neurodegenerative diagnoses. This demonstrates the clinical utility of a multiplex panel that can simultaneously capture both disease-specific and shared pathological signals. We built an ALS model including NEFL and other panel proteins, with remaining neurodegenerative diagnoses on the negative side.

### Limitations

This study was conducted at a single center with an internal train/test split, meaning performance may vary in other hospitals with different diagnostic mixes or blood-handling protocols. The PD and ALS groups were small, leaving operating points uncertain. The contrast group omitted key clinic differentials, such as multiple system atrophy for PD and cervical spondylotic myelopathy for ALS. Consequently, the models were not tested against the disorders that most often mimic these conditions in clinical practice.

## Conclusions

A single plasma neurodegeneration panel analyzed with machine learning can classify AD, PD, and ALS on the same assay. Multicenter external validation is required before clinical application, but this study suggests the Olink Target 48 Neurodegeneration panel has potential as a diagnostic tool for neurological clinics.

## Data Sharing Statement

Deidentified individual-participant data underlying this study are available from the corresponding author upon reasonable request, subject to institutional review and ethics approvals. Analysis scripts for the disease-specific Elastic Net classifiers are available at https://doi.org/10.5281/zenodo.22121272.

**eFigure 1.** Pearson correlation between Olink Target 48 Neurodegeneration NPX values and orthogonal assay measurements (SIMOA or HISCL). (A) pTau217 (SIMOA; r = 0.919; 95% CI, 0.897-0.937; n = 238); (B) GFAP (SIMOA; r = 0.898; 95% CI, 0.871-0.920; n = 238); (C) NEFL (SIMOA; r = 0.876; 95% CI, 0.842-0.903; n = 234); (D) Aβ40 (HISCL; r = 0.834; 95% CI, 0.791-0.869; n = 237); (E) Aβ42 (HISCL; r = 0.467; 95% CI, 0.361-0.561; n = 238); (F) Aβ42/Aβ40 (HISCL; r = 0.366; 95% CI, 0.251-0.471; n = 238). Each point represents one participant. The horizontal axis indicates the orthogonal assay measurement, and the vertical axis indicates the Olink Target 48 Neurodegeneration NPX value. Ordinary least-squares regression lines with 95% confidence bands are shown.

**eFigure 2.** Pearson correlation between High-Throughput (HT) and Olink Target 48 Neurodegeneration NPX values for 35 matched analytes comprising the full analytical cohort. The horizontal and vertical axes display HT and Olink Target 48 Neurodegeneration NPX values, respectively. Ordinary least-squares regression lines with 95% confidence bands are shown, and the Pearson *r* value is annotated on each panel. Each panel depicts a single matched analyte. Each point represents one participant.

**eFigure 3.** Principal-component analysis of normalized protein expression (NPX) profiles for 42 scaled proteins. (A) Score plot of PC1 versus PC2; (B) Score plot of PC1 versus PC3; (C) Score plot of PC2 versus PC3. Each point represents one participant, colored by diagnosis (HC, green; AD, red; PD, orange; ALS, blue; Others, purple). Axis labels indicate the percentage of variance explained by each component (PC1 17.2%, PC2 8.0%, PC3 6.4%; cumulative PC1-PC3 31.5%). Proteins were standardized before PCA. Score plots include 95% confidence ellipses for each diagnostic group.

**eFigure 4.** Heatmap of z-scored NPX values for 12 core assays (n=302). Rows represent individuals and columns represent assays. Samples are annotated by diagnostic group: HC, AD, PD, ALS, and others. Row annotations are colored by diagnosis (HC, green; AD, orange; PD, blue; ALS, red; Others, purple). Cell colors indicate full-cohort z-scored NPX values (scale −2 to 2), with higher values denoting increased abundance. Hierarchical clustering was performed using Ward linkage with Euclidean distance. The 12 assays included Aβ40, Aβ42, DDC, VSNL1, TREM2, GFAP, pTau217, EIF2AK2, NEFL, ENO2, S100B, and LRRK2.

**eFigure 5.** Plasma protein levels across diagnostic groups for 42 proteins. Samples are stratified by diagnosis: HC, AD, PD, ALS, and other conditions (Others). Each panel displays a box plot with individual data points representing single participants. Boxes and points are color-coded by diagnostic group (HC, blue; AD, magenta; PD, orange; ALS, red; Others, black). The vertical axis indicates normalized protein expression (NPX). Statistical significance was assessed using one-way ANOVA with Tukey post-hoc pairwise contrasts. Panels are arranged in ascending order of Benjamini-Hochberg-adjusted ANOVA P values. Brackets denote significant Tukey pairwise contrasts, indicated by asterisks; non-significant pairs are omitted.

**eTable 1.** Baseline characteristics of the Discovery training set (n = 211). Column definitions, group glosses, and reporting conventions are as in Table 1. The training set was obtained by diagnosis-stratified splitting of the analytical cohort (see Figure 1).

**eTable 2.** Baseline characteristics of the Internal validation testing set (n = 91). Column definitions, group glosses, and reporting conventions are as in Table 1. The testing set was held out for locked Train-Youden evaluation of disease classification models (see Figure 1).

## Additional Contributions

The authors would like to thank Ms. Natsumi Suzuki, Ayaka Morimoto, Azusa Oosumi, of the Department of Neuropsychiatry, Mr. Takayuki Hoshino, Ms. Yuka Hoshino, for their help with the PET examinations and image processing at Keio University School of Medicine.

