## Supplementary material for "Plasma Proteomic Classification of Alzheimer Disease, Parkinson Disease, and Amyotrophic Lateral Sclerosis Using a Single Neurodegeneration Panel: A Diagnostic Cohort Study": suplementary method, table 1-2 and figure 1-5

### eMethods

This eMethods file provides additional detail supporting the Methods in the main article. Primary design, eligibility, the 3 binary Elastic Net classifiers, and the locked Youden operating-point rule are summarized in the print Methods. Institutional approvals, registrations, and consent procedures are reported in full here.

Predicted class probabilities were obtained from the fitted logistic model:

$$\mathrm{logit}(p) = \beta_0 + \sum_j \beta_j x_j$$

$$p = \frac{1}{1 + e^{-\mathrm{logit}(p)}}$$

where $x_j$ denotes the preprocessed predictors retained with non-zero Elastic Net coefficients (continuous inputs were z-score standardized). Discrimination was summarized by ROC AUC, with a 95% confidence interval from percentile bootstrap resampling of hold-out predictions

#### Supplementary Table 1. Characteristics of the training set

| Characteristic | HC | AD | PD | ALS | Others | Total |
|---|---|---|---|---|---|---|
| Sample size, n | 44 | 65 | 27 | 19 | 56 | 211 |
| Male, n (%) | 27 (61.4) | 34 (52.3) | 18 (66.7) | 14 (73.7) | 29 (51.8) | 122 (57.8) |
| Female, n (%) | 17 (38.6) | 31 (47.7) | 9 (33.3) | 5 (26.3) | 27 (48.2) | 89 (42.2) |
| Age, mean ± SD | 68.1 ± 8.0 | 71.6 ± 9.6 | 70.5 ± 10.6 | 61.9 ± 8.8 | 69.1 ± 10.5 | 69.2 ± 9.9 |
| <50, n (%) | 0 (0.0) | 1 (1.5) | 2 (7.4) | 1 (5.3) | 2 (3.6) | 6 (2.8) |
| 50–64, n (%) | 16 (36.4) | 15 (23.1) | 4 (14.8) | 12 (63.2) | 13 (23.2) | 60 (28.4) |
| ≥65, n (%) | 28 (63.6) | 49 (75.4) | 21 (77.8) | 6 (31.6) | 41 (73.2) | 145 (68.7) |
| MMSE, mean ± SD | 29.0 ± 1.1 | 23.0 ± 6.0 | 27.6 ± 2.5 | — | 24.6 ± 6.3 | 25.4 ± 5.6 |
| CDR, mean ± SD | 0.0 ± 0.0 | 0.6 ± 0.4 | — | — | 0.5 ± 0.4 | 0.4 ± 0.4 |
| Hoehn and Yahr scale, mean ± SD | — | — | 2.5 ± 0.9 | — | — | 2.5 ± 0.9 |
| UPDRS-Ⅲ, mean ± SD | — | — | 34.6 ± 19.8 | — | — | 34.6 ± 19.8 |
| LEDD, mean ± SD | — | — | 395.5 ± 203.3 | — | — | 395.5 ± 203.3 |
| ALSFRS-R, mean ± SD | — | — | — | 39.6 ± 6.0 | — | 39.6 ± 6.0 |

#### Supplementary Table 2. Characteristics of the test set

| Characteristic | HC | AD | PD | ALS | Others | Total |
|---|---|---|---|---|---|---|
| Sample size, n | 18 | 28 | 12 | 8 | 25 | 91 |
| Male, n (%) | 9 (50.0) | 13 (46.4) | 7 (58.3) | 4 (50.0) | 11 (44.0) | 44 (48.4) |
| Female, n (%) | 9 (50.0) | 15 (53.6) | 5 (41.7) | 4 (50.0) | 14 (56.0) | 47 (51.6) |
| Age, mean ± SD | 69.0 ± 7.9 | 73.1 ± 9.7 | 67.2 ± 11.8 | 68.9 ± 12.6 | 72.6 ± 8.3 | 71.0 ± 9.6 |
| <50, n (%) | 0 (0.0) | 1 (3.6) | 0 (0.0) | 1 (12.5) | 0 (0.0) | 2 (2.2) |
| 50–64, n (%) | 6 (33.3) | 4 (14.3) | 6 (50.0) | 0 (0.0) | 5 (20.0) | 21 (23.1) |
| ≥65, n (%) | 12 (66.7) | 23 (82.1) | 6 (50.0) | 7 (87.5) | 20 (80.0) | 68 (74.7) |
| MMSE, mean ± SD | 28.8 ± 0.9 | 23.5 ± 5.0 | 27.5 ± 2.2 | — | 25.5 ± 4.8 | 25.8 ± 4.5 |
| CDR, mean ± SD | 0.0 ± 0.0 | 0.5 ± 0.3 | — | — | 0.6 ± 0.6 | 0.4 ± 0.5 |
| Hoehn and Yahr scale, mean ± SD | — | — | 2.5 ± 0.5 | — | — | 2.5 ± 0.5 |
| UPDRS-Ⅲ, mean ± SD | — | — | 31.2 ± 10.5 | — | — | 31.2 ± 10.5 |
| LEDD, mean ± SD | — | — | 491.4 ± 260.9 | — | — | 491.4 ± 260.9 |
| ALSFRS-R, mean ± SD | — | — | — | 38.4 ± 10.4 | — | 38.4 ± 10.4 |

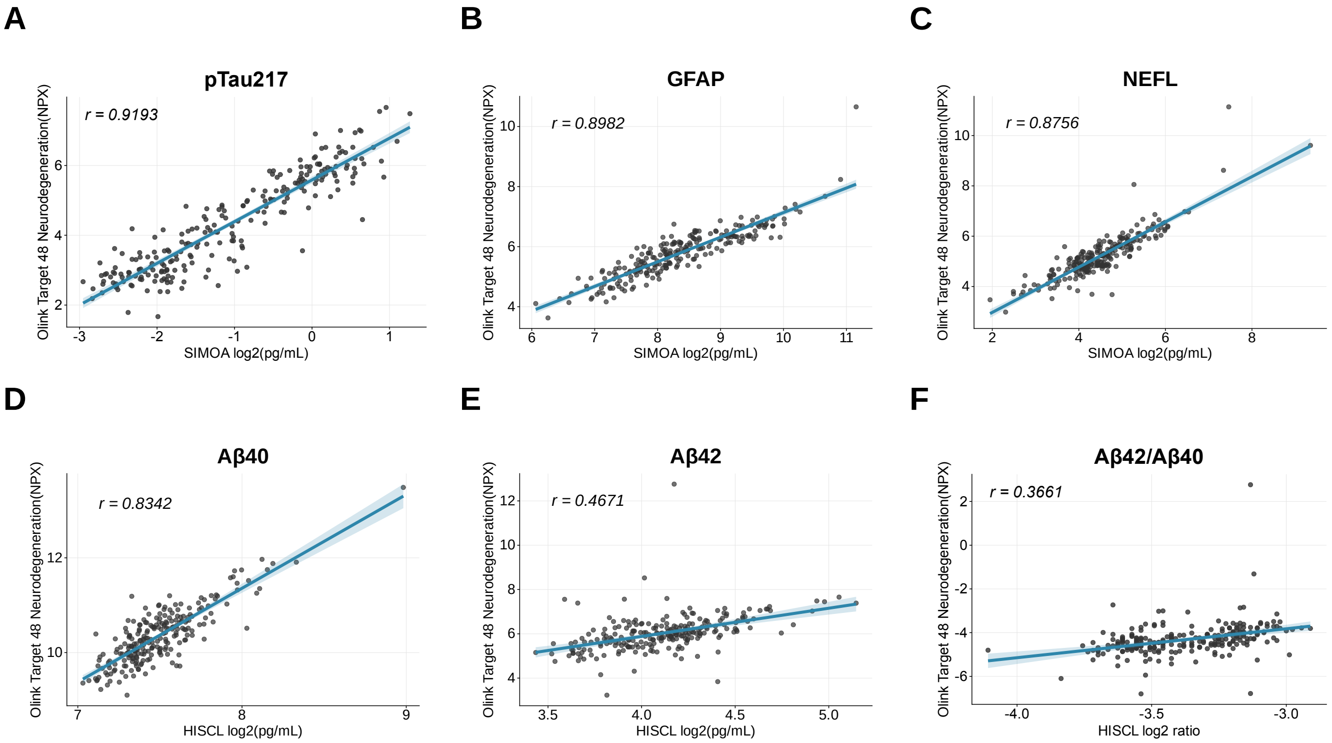
A
pTau217
r = 0.9193
Olink Target 48 Neurodegeneration(NPX)
SIMOA log2(pg/mL)
B
GFAP
r = 0.8982
Olink Target 48 Neurodegeneration(NPX)
SIMOA log2(pg/mL)
C
NEFL
r = 0.8756
Olink Target 48 Neurodegeneration(NPX)
SIMOA log2(pg/mL)
D
Aβ40
r = 0.8342
Olink Target 48 Neurodegeneration(NPX)
HISCL log2(pg/mL)
E
Aβ42
r = 0.4671
Olink Target 48 Neurodegeneration(NPX)
HISCL log2(pg/mL)
F
Aβ42/Aβ40
r = 0.3661
Olink Target 48 Neurodegeneration(NPX)
HISCL log2 ratio

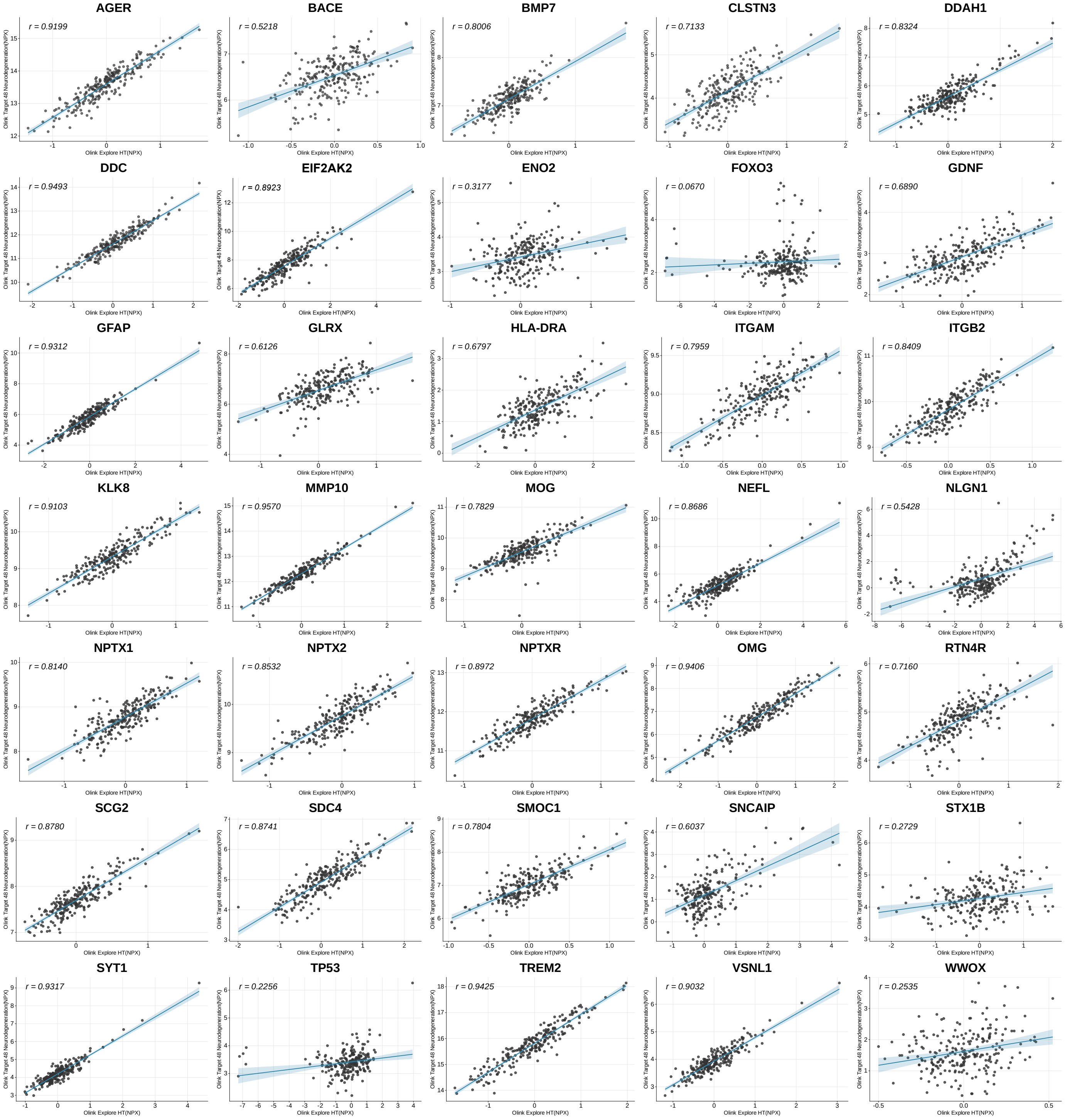
AGER
r = 0.9199
BACE
r = 0.5218
BMP7
r = 0.8006
CLSTN3
r = 0.7133
DDAH1
r = 0.8324
DDC
r = 0.9493
EIF2AK2
r = 0.8923
ENO2
r = 0.3177
FOXO3
r = 0.0670
GDNF
r = 0.6890
GFAP
r = 0.9312
GLRX
r = 0.6126
HLA-DRA
r = 0.6797
ITGAM
r = 0.7959
ITGB2
r = 0.8409
KLK8
r = 0.9103
MMP10
r = 0.9570
MOG
r = 0.7829
NEFL
r = 0.8686
NLGN1
r = 0.5428
NPTX1
r = 0.8140
NPTX2
r = 0.8532
NPTXR
r = 0.8972
OMG
r = 0.9406
RTN4R
r = 0.7160
SCG2
r = 0.8780
SDC4
r = 0.8741
SMOC1
r = 0.7804
SNCAIP
r = 0.6037
STX1B
r = 0.2729
SYT1
r = 0.9317
TP53
r = 0.2256
TREM2
r = 0.9425
VSNL1
r = 0.9032
WWOX
r = 0.2535
Olink Target 48 Neurodegeneration(NPX)
Olink Explore HT(NPX)

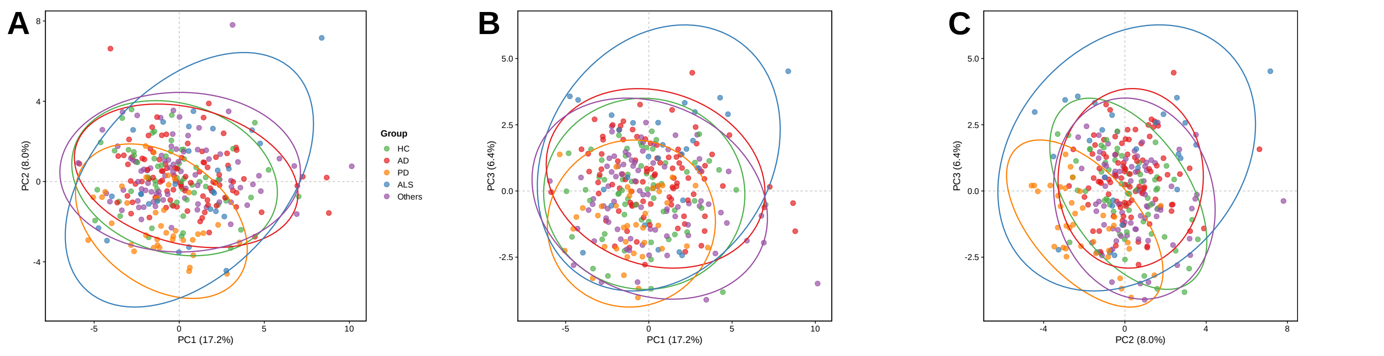
A
B
C
PC1 (17.2%)
PC2 (8.0%)
PC3 (6.4%)
Group
HC
AD
PD
ALS
Others

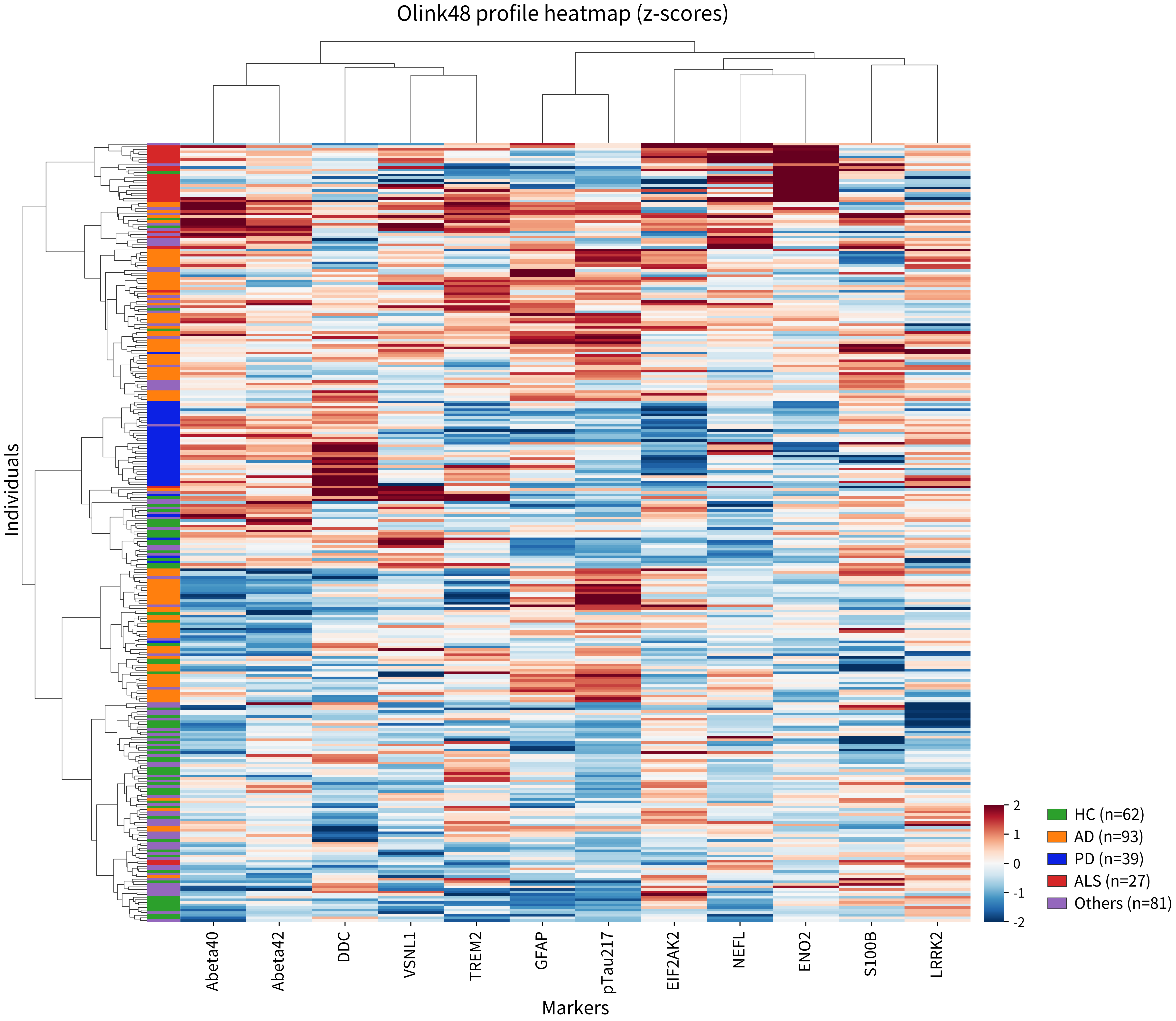

Olink48 profile heatmap (z-scores)
Individuals
Markers
Abeta40
Abeta42
DDC
VSNL1
TREM2
GFAP
pTau217
EIF2AK2
NEFL
ENO2
S100B
LRRK2
2
1
0
-1
-2
HC (n=62)
AD (n=93)
PD (n=39)
ALS (n=27)
Others (n=81)

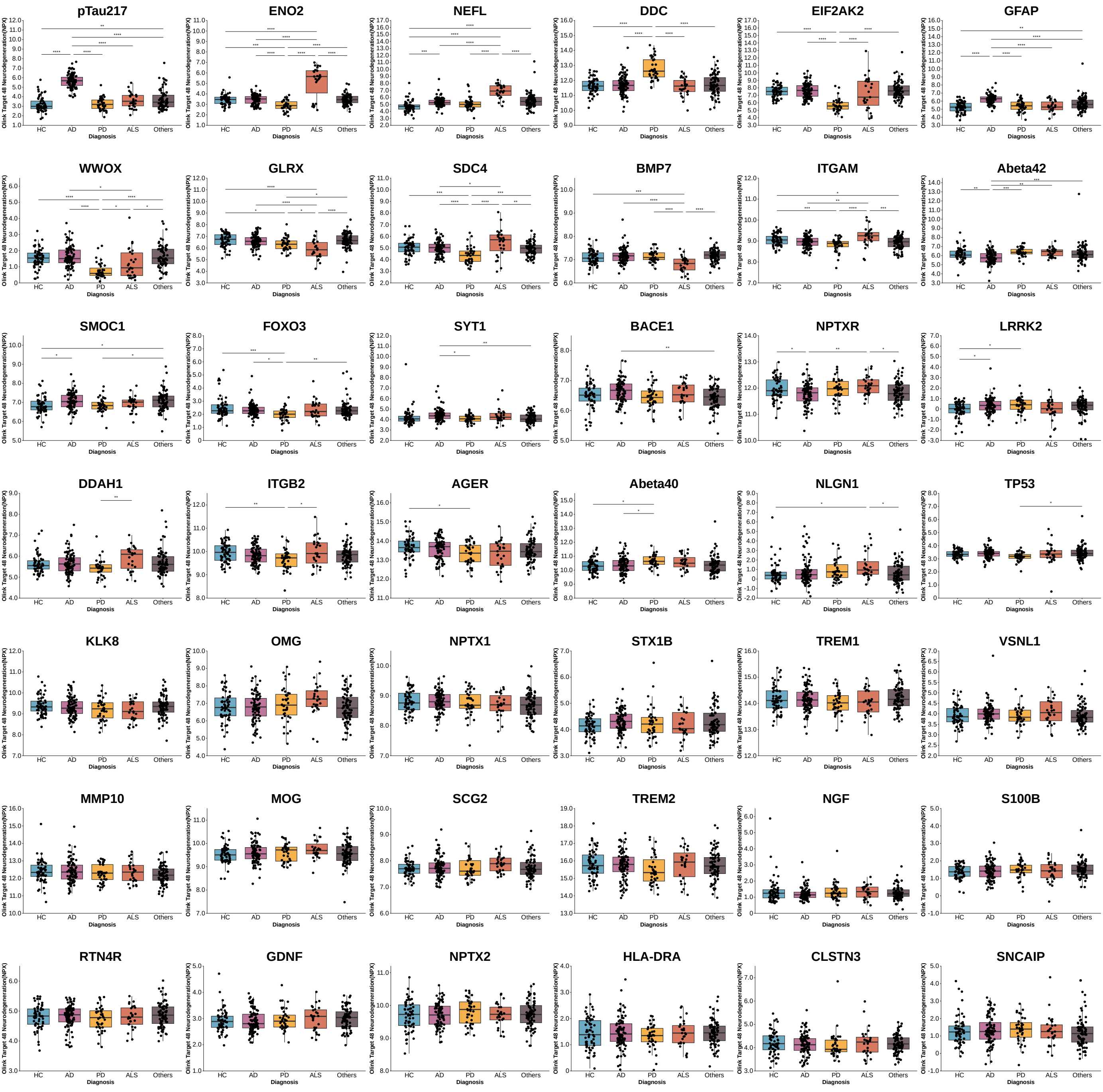

pTau217
ENO2
NEFL
DDC
EIF2AK2
GFAP
WWOX
GLRX
SDC4
BMP7
ITGAM
Abeta42
SMOC1
FOXO3
SYT1
BACE1
NPTXR
LRRK2
DDAH1
ITGB2
AGER
Abeta40
NLGN1
TP53
KLK8
OMG
NPTX1
STX1B
TREM1
VSNL1
MMP10
MOG
SCG2
TREM2
NGF
S100B
RTN4R
GDNF
NPTX2
HLA-DRA
CLSTN3
SNCAIP
Olink Target 48 Neurodegeneration(NPX)
HC
AD
PD
ALS
Others
Diagnosis
